# Factors associated with self-reported health among New Zealand military Veterans: a cross-sectional study

**DOI:** 10.1101/2021.08.22.21262319

**Authors:** David McBride, Ari Samaranayaka, Amy Richardson, Dianne Gardner, Emma Wyeth, Brandon De Graaf, Sarah Derrett

## Abstract

**Objective:** To identify factors associated with better or poorer self-reported health status in New Zealand Military Veterans.

**Design:** An online cross-sectional survey.

**Participants:** The total number of eligible Veterans is unknown, but a total of 1,817 Veterans responded, including 1009 serving personnel providing a 26% response rate from that group.

**Study variables:** Health status was self-reported using the EQ-5D-5L, which asks about problems across five dimensions (mobility, self-care, usual activities, pain/discomfort, and anxiety/depression), with five levels of severity (e.g. no, slight, moderate, severe or extreme problems). The EQ-5D-5L also contains a visual analogue scale (EQ-VAS), scaled from 0 (worst) to 100 (best) imagined health. Hypothetical relationships with better health were positive social support, sleep and psychological flexibility; with poorer health, exposure to psychological trauma, distress and hazardous drinking.

**Results:** The proportion of Veterans reporting ‘any problems’ compared to ‘no problems’ with the five EQ-5D dimensions, was similar to those found in the general NZ population, although a higher proportion of Veterans reported problems with mobility, self-care, usual activities and pain/discomfort. Psychological flexibility and better sleep quality were associated with higher EQ-VAS scores; distress was associated with lower EQ-VAS scores.

**Conclusion:** In this sample of New Zealand Veterans, psychological flexibility and good sleep are associated with better self-rated health, and distress and poor sleep with diminished health. These factors might be used as sentinel health indicators in assessing Veteran health status. As distress, psychological flexibility and sleep are closely related, cognitive behavioural therapy encompassing these domains may be useful in improving the health of New Zealand Veterans.

**Strengths and limitations of this study:**

- Many studies of Veterans have focused on adverse outcomes, but we have been able to focus on a holistic measure of ‘health’.
- The study was sufficiently powered to detect important relationships indicating opportunities for intervention.
- The exact response rate is unknown, and possible bias may be a limitation.
- The cross-sectional design means that we cannot explore cause and effect relationships.

## INTRODUCTION

The three major events in the military life course are entry to military service, deployment on active service and transition back to civilian life. On entry, soldiers, sailors and air personnel are subject to a selection process to ensure, as far as possible, good physical and mental health, giving rise to the ‘healthy soldier effect’, with service personnel being, on average, healthier than the general population.[1] However, the physical and psychological stressors of military service have been found to erode this effect.[2]

The physical stressors have a particular impact on the lower limb, with load carriage, high intensity training and the design of footwear being implicated in injury causation.[3] A military career increases opportunities for psychological trauma, and post-traumatic stress disorder (PTSD) has been identified as the ‘signature injury’ of United States service men and women deployed to Afghanistan and Iraq.[4]

A focus on adverse health events in the literature[5] means that wellbeing is relatively overlooked. In the long run, military service has been found to have positive effects.[6,7] Good health after service does, however, depend on the success of the ‘military civilian transition’, a complex process for which models have been developed.[8] Health problems developed in service, difficulty in assuming a post-service identity and many other factors contribute to health and wellbeing outcomes.[8] Providing health support to New Zealand Veterans is the responsibility of New Zealand Veteran’s Affairs, however a Veteran is only eligible for support if they have deployed on legally defined ‘qualifying operational service.’[9] The total number of Veterans is unknown, however there are few Second World War Veterans left, and the Malayan, Korean, Borneo and Vietnam Veterans, a total of approximately 12,000-13,000 personnel, form a significant proportion of the Veterans supported by NZVA.[10] The only comprehensive follow up has been on the health of the 3,394 Vietnam Veterans.[11] The next major deployment was that to Bosnia in 1992, followed by a series of ‘Company Group’ deployments, combat troops with support elements, of 200-350 personnel each. culminating with the withdrawal of the Provincial Reconstruction Team from Bamian, Afghanistan, in 2019, a total of approximately 10,000 Veterans. There have in addition been approximately 10,000 Veterans deployed on United Nations duties, also on ships and aircraft.

The aims of this study were to describe self-reported health among these more recently deployed New Zealand Veterans, and identify factors associated with better or poorer health.

### Veteran and public involvement

At the design stage, we sought advice on design from the Ministerial Veterans Health Advisory Panel, also forming a steering group with representatives from the New Zealand Defence Force, New Zealand Veterans Affairs, the Royal New Zealand Returned and Services Association (RSA)[12] and No Duff,[13] a charity providing an urgent support capability to Veterans and their families. We also consulted with the Ngāi Tahu Research Consultation Committee in order to assess the importance of the project to Māori, New Zealand’s indigenous population. We undertook to inform the Veteran community before releasing the results.

## METHODS

### Participants

Data were collected via an online survey; a posted hard copy version was also available on request. There is no comprehensive Veteran registry, however In June 2018, a link to the online questionnaire was sent by email to all currently serving regular and reserve New Zealand Defence Force (NZDF) members registered on the NZDF email system who had qualifying operational service, as indicated by holding the New Zealand Operational Service Medal, numbering 3,874 personnel at that time. An introductory message and link to the questionnaire were also presented on the NZDF ‘intranet landing page’, a secure internal webpage from which all regular force personnel can access relevant work-related content, tools, and resources. The steering group recommended that no restriction be placed on participation, so retired military personnel were invited to participate through posters distributed to reserve units and the 43 local social clubs identified by the RSA national office to be ‘Veteran active’. Paper questionnaires with return postage envelopes were made available at these sites. Announcements were also made on military social media pages, and both retired and currently serving personnel were invited to participate through an announcement on the No Duff website. The questionnaire was available for completion from June to December 2018. Ethical approval for the study was obtained from the Northern B Health and Disability Ethics Committee, reference 17NTB118.

### Questionnaire

#### Criterion variable

Self-rated health status was assessed using the EQ-5D-5L,[14] a short questionnaire asking about the respondent’s health across five dimensions: mobility, self-care, usual activities, pain/discomfort and anxiety/depression, with response options ranging from (e.g.) ‘no problems’, to ‘extreme problems’. For each dimension, participants were categorised as having ‘any problems’ if they selected any response other than ‘no problems’.

Additionally, the EQ-5D-5L visual analogue scale (EQ-VAS) asks the respondent to mark on a vertical visual analogue scale (VAS) how good or bad their health is today, where the endpoints are labelled ‘the best health you can imagine’ (score of 100) and ‘the worst health you can imagine’ (score of 0).

#### Independent variables

Demographic characteristics included age, sex, ethnicity, service years, and past deployment (yes/no). Ethnicity prioritisation was adopted,[15] whereby participants with multiple responses were assigned to one of the categories, in the order of Māori, Pacific Peoples, Other and European.

General psychiatric morbidity was assessed using the 12-item General Health Questionnaire (GHQ-12),[16] scored using a four point scale (0-3) and summing the 12 items to give a total score, with higher scores indicating elevated distress.

Social support was measured using the Social Provisions Scale,[17] with responses made on a four-point Likert-type scale ranging from 1 ‘strongly disagree’ to 4 ‘strongly agree’. The 24 items can be reduced to six subscales (attachment, social integration, reassurance of worth, reliable alliance, social guidance, opportunity for nurturance) or summed to create a total score, with greater scores indicating greater social support.

Alcohol use was measured using the AUDIT-C,[18] scaled from 0-12. A score of 3+ for women and 4+ for men indicated potentially hazardous drinking behaviour.

Sleep quality was assessed with the Sleep Condition Indicator (SCI), [19] assessing insomnia as described in the Diagnostic and Statistical Manual of Mental Disorders version 5 (DSM-V).[20] The SCI consists of eight items rated from 0-4, the total scores being scaled to a range of 0 to 10, where higher scores represent better sleep.

Trauma exposure was assessed with the Brief Trauma Scale (BTS),[21] which captures past exposure to situations that were life threating or capable of producing serious injury.

Psychological flexibility was measured with the 10-item AAQ-II, designed as a measure of effectiveness in a particular mode of behavioural intervention, Action and Commitment Therapy (ACT).[22] Items were answered on a 7-point scale, with options ranging from ‘never true’ to ‘always true’. The items were summed to obtain a total score (possible range 10 to 70), with higher scores indicative of greater psychological flexibility.

### Statistical analyses

With respect to the calculation of summed scores, if only one item was missing for a particular measure then this was imputed with the mean of the remaining items; if more than one item was missing then the score was set to missing for that participant. Complete case analysis was used in the remaining analyses. The five dimensions of the EQ-5D-5L were compared to the NZ population normative values.[19]

Univariate ordinary least-squares linear regression analyses assessed the strength of relationships between each independent variable and EQ-VAS scores, using robust standard errors to account for heteroscedasticity and calculating 95% confidence intervals (95% CIs). Multivariable linear regression was then used to identify the role of the independent variables while adjusting for each other. None of the social support sub-scales were used in this multivariable model, instead using the social support total score. The model was built using backward variable selection with p<0.10 for variable retention, with the exceptions of age, sex, service years, and deployment status which were retained as adjusting variables irrespective of p-values.

## RESULTS

When the survey went online, invitations were emailed to the 3784 serving Veterans in the NZDF, resulting in 1009 responses, 26% of that group, added to by 449 retired and 288 non-deployed Veterans. a total of 1817, 90 of whom completed the paper questionnaire, 1767 (97%) completing the EQ-VAS and were thus included in all the analyses.

A Supplementary table presents the EQ-VAS score according to the sample characteristics. Figure 1 shows the proportion of EQ-5D-5L dimension responses reporting ‘any problem’ severity level other than ‘no problems’ in comparison to the New Zealand population normative values.[23]

**Figure 1.**
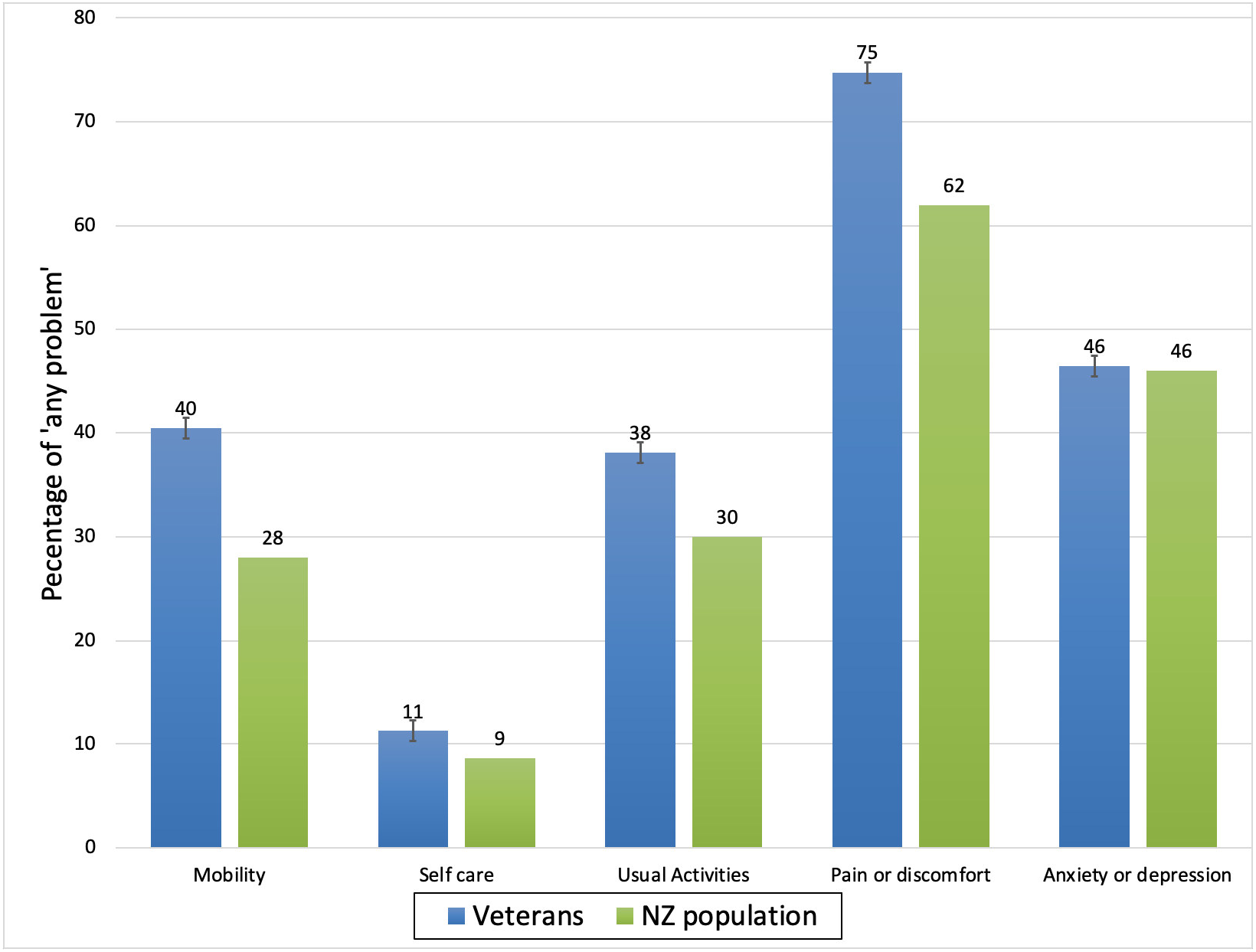
The proportion of Veterans reporting ‘any problem’ with each of the EQ-5D-5L dimension scores compared to the NZ population normative proportions.

The results of the univariate analysis are displayed in Table 1. Here, for continuous characteristics, the regression coefficient (*ß*) represents the change in the mean EQ-VAS associated with one unit increase in the characteristic. For categorical characteristics, *ß* is the change in mean EQ-VAS scores compared to the referent category.

**Table 1:** Univariate cross-sectional associations between variables and mean EQ-VAS scores for New Zealand Veterans (N=)

| Characteristic | *n* | $\beta$ | 95% CIs | *p*-value |
| --- | --- | --- | --- | --- |
| **Age\*** | 1762 | -0.19 | [-0.24, -0.13] | <.01 |
| **Sex** |  |  |  |  |
| Female | 220 | Reference | - |  |
| Male | 1520 | -0.65 | [-3.04, 1.73] | .59 |
| **Ethnicity** |  |  |  |  |
| NZ European | 1382 | Reference | - |  |
| Māori | 245 | 1.21 | [-1.00, 3.42] |  |
| Other | 140 | -2.95 | [-6.26, 0.36] | 0.10 |
| **Service years\*** | 1670 | 0.19 | [0.11, 0.27] | <.01 |
| **Deployment status** |  |  |  |  |
| Not deployed | 288 | Reference | - |  |
| Deployed | 1458 | 5.80 | [ 3.35, 8.25 ] | <.01 |
| **GHQ-12 score\*** | 1765 | -1.63 | [-1.79, -1.47] | <.01 |
| **Social support\*** |  |  |  |  |
| Attachment | 1760 | 2.02 | [ 1.69, 2.34 ] | <.01 |
| Social integration | 1758 | 2.45 | [ 2.06, 2.88 ] | <.01 |
| Reassurance of worth | 1756 | 2.48 | [ 2.11, 2.85 ] | <.01 |
| Reliable alliance | 1760 | 2.30 | [ 1.91, 2.70 ] | <.01 |
| Social guidance | 1760 | 2.00 | [ 1.65, 2.35 ] | <.01 |
| Opportunity for nurturance | 1758 | 0.85 | [ 0.47, 1.23 ] | <.01 |
| Social support total score | 1753 | 0.52 | [ 0.44, 0.59 ] | <.01 |
| **Psychological flexibility\*** | 1750 | 0.79 | [ 0.71, 0.87 ] | <.01 |
| **Sleep score\*** | 1747 | 3.47 | [ 3.11, 3.83 ] | <.01 |
| **AUDIT-C** | 1691 |  |  |  |
| Non-hazardous |  | Reference | - |  |
| Hazardous |  | 0.62 | [-1.03, 2.26] | 0.46 |
| **Exposure to traumatic events** | 1754 |  |  |  |
| Not exposed |  | Reference | - |  |
| Exposed |  | -5.72 | [-7.34, -4.11] | <.01 |

\*Scored as continuous variables, coefficient is per unit increase.

Of the demographic variables, age was associated with lower EQ-VAS scores, length of service with higher scores; no relationships were apparent for sex or ethnicity. Positive coefficients, indicating better health with presence of the characteristic, were present for dichotomous variables of deployment, where the mean EQ-VAS score for deployed Veterans was 5.8 VAS units higher (better) than for non-deployed. The largest negative associations were for exposure to traumatic events, with a mean EQ-VAS score 5.7, lower for those exposed compared to those not exposed. Distress, as measured by the GHQ-12, had a negative association with health state. Positive associations with health state were found for psychological flexibility as measured by the AAQ-II, better sleep scores, and most of the dimensions of social support, barring ‘opportunity for nurturance’.

Table 2 shows results from two models. The first model is adjusted for the other characteristics, with 11 variables and 1,557 people providing valid responses for all factors included in the model. All effect sizes were reduced, and the social support and AUDIT-C scores were no longer associated, with 11 variables explaining 35% of the variability in the EQ-VAS.

**Table 2.** Multivariable models of associations between variables and mean EQ-VAS scores for New Zealand Veterans.

| Characteristic | Adjusted model, N used =1,557 |  |  | Final model, N used =1,600 |  |  |
| --- | --- | --- | --- | --- | --- | --- |
| | $\beta$ | 95% CI | p-value | $\beta$ | 95% CI | p-value |
| **Age (years)\*** | 0.17 | [-0.23, 1.11] | <0.01 | -0.17 | [-0.23, -0.12] | <0.01 |
| **Sex** |  |  |  |  |  |  |
| Female | Ref | - | - | Ref | - | - |
| Male | 0.24 | [-2.52, 2.03] | 0.84 | -0.60 | [-2.82, 1.63] | 0.60 |
| **Ethnicity** |  |  |  |  |  |  |
| NZ European | Ref | - | - | Ref | - | - |
| Māori | 0.29 | [-1.70, 2.27] | 0.07 | 0.19 | [-1.77, 2.14] |  |
| Other | -3.58 | [-6.66, -0.51] |  | -2.91 | [-5.88, 0.06] | 0.15 |
| **Service years\*** | 0.17 | [ 0.10, 0.25] | <0.01 | 0.17 | [ 0.10, 0.25] | <0.01 |
| **Deployment status** |  |  |  |  |  |  |
| Not deployed | Ref |  |  | Ref | - |  |
| Deployed | 2.91 | [ 0.64, 5.18] | <0.05 | 2.90 | [0.65, 5.15] | 0.01 |
| **GHQ12 score\*** | -0.87 | [-1.09, -0.65] | <0.01 | -0.92 | [-1.13, -0.71] | <0.01 |
| **Social support (SPS score)\*** | 0.01 | [-0.07, 0.10] | 0.76 | - | - | - |
| **Psychological flexibility score (AAQii)\*** | 0.26 | [ 0.15, 0.37] | <0.01 | 0.24 | [0.14, 0.35] | <0.01 |
| **Sleep score (SCI), Range 0-10\*** | 1.60 | [ 1.18, 2.02] | <0.01 | 1.63 | [1.22, 2.05] | <0.01 |
| **AUDIT_C score** |  |  |  |  |  |  |
| Non hazardous | Ref |  |  | - | - | - |
| Hazardous | 0.41 | [-1.00, 1.81] | 0.57 | - | - | - |
| **Exposure to traumatic events (BTQ)** |  |  |  |  |  |  |
| Not exposed | Ref |  |  | Ref | - | - |
| Exposed | -1.77 | [-3.30, -0.24] | <0.05 | -1.81 | [-3.32, 0.30] | 0.02 |
| | | | $R^2 = 0.35$ | | | $R^2 = 0.36$ |

\*Scored as continuous variables, coefficient is per unit increase.

The final model involved backward variable selection setting a p-value of 0.10, identifying a smaller subset of variables. Age, sex, service years, and deployment status were retained in the model irrespective of their p-values, thus adjusting for those variables, using 1,600 complete responses. Social support and AUDIT-C hazardous drinking were not retained in this final model; other coefficients remaining essentially the same, with a minimal effect on the overall *R*^2^.

## DISCUSSION

### Principal findings

In general, Veterans had a similar proportion of ‘any problem’ responses in the EQ-5D dimensions as the general population in New Zealand, with evidence of more problems in the physical domains of mobility, usual activities and pain and discomfort, but no difference in the psychological domain.

Mutual adjustment, with 11 variables in the model, reduced all the effect sizes and explained 35% of the variance, thus leaving 65% which cannot be explained. The final model had 9 variables, explaining 36% of the variance. The results make conceptual sense in that distress is associated with reduced EQ-VAS, while psychological flexibility is associated with a modest protective effect. Surprisingly, social support was not identified as an associated factor, however we may not have measured some support domains valued by Veterans. There are known to be many other influences on health, including ‘social wellbeing’,[5] financial status, personality and non-deployment related stressors,[25] which we have not measured.

### Strengths and weaknesses

Strengths of our study were the relatively large sample size, the inclusion of all Veterans, the assessment of ‘health’, infrequently investigated in Veteran populations, and the inclusion of New Zealand Veterans with a range of characteristics, including ‘deployed’ and ‘non-deployed’ Veterans. As a measure of health, the EQ-5D-5L dimensions and EQ-VAS ask about health on the day that respondents complete the questionnaire, the EQ-VAS end points being, respectively, the ‘best’ and ‘worst’ health they can imagine, so it is a holistic measure of health state.[24]

The response rate of 27% from serving Veterans, along with the unknown total number of Veterans, raises the question of bias, the direction of which is difficult to assess, as responses may be more likely from either Veterans with good or poor health. There are also likely to be other personal characteristics that we have not measured. The cross-sectional design also means that we cannot explore cause and effect, so recommendations for future interventions require additional support from longitudinal studies.

### Comparison with other studies

We have previously reported factors associated with post-traumatic stress in this group,[26] using the Military Post Traumatic Stress Checklist (PCL-M). Factors associated with higher PCL-M scores were trauma exposure, older age, male gender, and being of Māori ethnicity. Factors associated with lower PCL-M scores were greater length of service, psychological flexibility, and better quality sleep. Using health as the outcome disclosed that Māori did not have poorer self-reported health compared to non-Māori, that deployment had a positive effect, and in the univariate models, all of the dimensions of social support were associated with improved health. The final model also included good sleep and psychological flexibility, providing most of the explanatory power in the model.

No other studies appear to have used the EQ-VAS as an outcome measure for Veteran health. Boehmer et al.[27] examined wellbeing among participants in the 2000 Behavioural Risk Factor Surveillance System describing health-related quality of life (HRQoL) by sex and military status, active duty, reservists Veterans, or no military service. Participants were asked to rate recent physical health, mental health, and activity limitation. Active duty men were more likely than men without military service to report 14 or more days of activity limitation, pain, and not enough rest in the past 30 days. Reserve personnel reported better overall HRQoL than non-military participants, and no difference was observed between Veterans and persons with no military service. There are also reports indicating that non-deployed personnel retain better health than those who have been deployed.[28] Notably, the predominant reason for medical discharge from the British Armed Forces was musculoskeletal problems.[29]

Diaz Santana et al.[30] carried out a cross-sectional survey of 60,000 U.S. Veterans of Afghanistan and Iraq, with 20,563 responses. Mental quality of life scores were higher among the non-deployed group compared to the deployed group, though the deployed group reported higher physical quality of life scores compared to the non-deployed. Both mental and physical quality of life were lower among Veterans compared to U.S. population norms. Both positive and negative consequences of deployment have been described.[7,31] In a study of Dutch Veterans[7], two out of three reported a positive effect of deployment on their quality of life at the time of the survey, this being related to positive feelings such as satisfaction or comradeship, but a few having emotions such as frustration or shame. As regards tangible effects[31], negative consequences included the military ‘chain of command’, being away from home, and deterioration of marital/significant other relationships. Positive influences include improved financial security, self-improvement, and time to reflect.

Sleep difficulties are a common symptom for those with PTSD. McCarthy et al.[32] reported on the 3,157 U.S. military Veterans enrolled in the National Health and Resilience in Veterans Study, in which 27.6% reported poor sleep quality. Path analyses revealed significant associations between poor sleep, severity of PTSD, poorer mental and physical health functioning and lower overall quality of life.

Most Veterans do cope well with a military career, and service has a positive effect on wellbeing. However, Oster et al.[5] emphasise that when things do not go well for Veterans, their mental, physical and social health is interconnected, so their needs can be complex, and management requires an integrated approach.

### Future directions

The results suggest that distress, psychological flexibility, and sleep have an important relationship with self-rated health among eterans in this study.

Reducing distress through the promotion of psychological flexibility might be possible, although our finding here must be subject to caution as several researchers argue that the AAQ-II may be measuring psychological distress and affect rather than psychological inflexibility.[33] Psychological flexibility is specifically targeted by ACT, a psychological intervention described as being in the ‘third wave’ of behaviour change strategies.[34] The six core processes of ACT (acceptance, cognitive defusion, being present, self as context, values, and committed action) aim to increase psychological flexibility, the goal being “to have clients experience the world more directly so that their behavior is more flexible and thus their actions more consistent with the values that they hold”.[34] Approaches such as ACT may therefore improve health state in appropriate subjects. Lang et al. carried out a randomised clinical trial (RCT), comparing ACT with person-centred therapy,[35] showing a general improvement in symptoms of distress across both treatment arms, ACT providing superior improvement in insomnia. The drop-out rate for both therapies was however high, and the two groups did not exhibit any change in psychological flexibility. The authors proposed that future studies should include additional measures of ACT processes to determine which are actually affected by ACT.

Sleep in military personnel has been recognised as a ‘vital health behaviour’ for which policies and guidelines must be developed.[36] Cognitive behavioural therapy for insomnia (CBT-I) is regarded as an effective ‘first line’ treatment, and a brief intervention has been described for use in Australian general practice.[37] The Lange et al trial[35] showed CBT-I to be effective, however future studies should include outcome measures that include ACT processes. Our final model showed that distress had a negative association with health, and psychological flexibility had a positive relationship, with sleep most likely related to both of these variables. It would seem important to screen for these conditions prior to transition from the military, as well as among retired Veterans, in order to provide targeted support. Further research is needed to examine the potential of ACT to improve Veterans’ wellbeing.

## Data Availability

Data from this study is unsuitable for public deposition due to the privacy of participant data. Data are anonymised, but contain information on deployments (including location and duration) which could lead to some participants being identified. Furthermore, the participant information sheet, as required by the Southern Health and Disability Ethics Committee specifically contains the statement that 'all study data would be kept strictly confidential to the research team,' Qualified researchers may apply for data access with the research team at and/or

## DATA SHARING STATEMENT

Data from this study is unsuitable for public deposition due to the privacy of participant data. Data are anonymised, but contain information on deployments (including location and duration), which could lead to some participants being identified. Furthermore, the participant information sheet, as required by the Southern Health and Disability Ethics Committee specifically contains the statement that ‘all study data would be kept strictly confidential to the research team.’ Qualified researchers may apply for data access with the research team at and/or.

## CONTRIBUTION

**Investigation:** Amy Richardson, Emma H. Wyeth, Sarah Derrett, Daniel Shepherd, David McBride.

**Methodology:** Amy Richardson, Ari Samaranayaka, Dianne Gardner, Emma H. Wyeth, Sarah Derrett, David McBride. Daniel Shepherd

**Project administration:** Amy Richardson

**Resources:** Brandon deGraaf

**Software:** Brandon deGraaf

**Supervision:** Ari Samaranayaka

**Validation:** Amy Richardson

**Visualization:** Amy Richardson

**Writing – original draft:** David McBride. AS, DG, EHW, SD and DS contributed to the re-writes and final draft

## ACKNOWLEDGEMENTS

We would like to thank Shane Harvey for his input into the initial design, Aidan Smith, previous research advisor to the NZDF, for steering us through the consents process, also the members of our steering group: COL Clare Bennett (NZDF); Ms Marti Eller (New Zealand Veterans Affairs); Mark Compain, Danny Nelson and Richard Terrill (RSA); and NoDuff representatives Aaron Wood and Lars Millar.

Finally, a thank you to the New Zealand Veterans who took the time and effort to complete a rather tedious questionnaire. Kia Kaha.

## REFERENCES

1. McLaughlin R, Nielsen L, Waller M. An Evaluation of the Effect of Military Service on Mortality: Quantifying the Healthy Soldier Effect. Ann Epidemiol 2008;18(2):928–936.

2. Bollinger MJ, Schmidt S, Pugh JA et al. Erosion of the healthy soldier effect in Veterans of US military service in Iraq and Afghanistan. Popul Health Metr. 2015;13:8.

3. Kimberley A. Andersen KA, Grimshaw PN et al. Musculoskeletal Lower Limb Injury Risk in Army Populations. Sports Med-Open 2016;2:22. DOI 10.1186/s40798-016-0046-z.

4. Hoge CW, Castro CA, Messer SC et al. Combat duty in Iraq and Afghanistan, mental health problems, and barriers to care. N Engl J Med 2004; 351(1):13–22.

5. Oster C, Morello A, Venning A et al. The health and wellbeing needs of Veterans: a rapid review. BMC Psychiatry. 2017;17(1):414.

6. Spiro A, 3rd, Settersten RA, Aldwin CM. Long-term Outcomes of Military Service in Aging and the Life Course: A Positive Re-envisioning. Gerontologist. 2016;56(1):5–13.

7. Duel J and Reijnen A. The long term effects of military deployment and their relation with the quality of life of Dutch Veterans. Mil Behav Heal. 2021;9(2):160–169.

8. Pedlar D, Thompson JM, Castro CA. Military Veteran reintegration. Military-to-civilian transition theories and frameworks. In: Castro C, Dursun S, editors. Military Veteran reintegration, approach, management, and assessment of military Veterans transitioning to civilian life. San Diego: Academic Press; 2019. 257p.

9. Veterans Support Act 2014. (NZ) Available from: http://www.legislation.govt.nz/act/public/2014/0056/60.0/DLM5537774.html.

10. New Zealand History [Internet]. Post Second World War. Available: https://nzhistory.govt.nz/war/post-second-world-war

11. New Zealand Veterans Affairs [Internet]. Research about New Zealand’s Vietnam Veterans. Available: https://www.Veteransaffairs.mil.nz/about-Veterans-affairs/our-documents-and-publications/research/research-about-new-zealands-viet-nam-Veterans/

12. Royal New Zealand Returned and Services Association [internet]. Available: https://www.rsa.org.nz/

13. NoDuff. [Internet]. Avialable: https://www.noduff.org/

14. EuroQol Research Foundation. EQ-5D-5L User Guide[Internet]. Rotterdam: EuroQol Research Foundation; 2021[cited 27th July 2020]. Available from: https://euroqol.org/eq-5d-instruments/eq-5d-5l-about/

15. Health Information Standards Organisation. Ethnicity data protocols. Wellington: Ministry of Health; 2017. Standard No.: HISO 10001:2017.

16. Goldberg D.P. The Detection of Psychiatric Illness by Questionnaire. Maudsley Monograph No. 21. London: Oxford University Press; 1972.

17. Barrera, M., Jr. and Ainlay, S.L. The structure of social support: a conceptual and empirical analysis. J Comm Psychol 1983;11(2):133–43.

18. Bush K, Kivlahan DR, McDonell MB et al. The AUDIT alcohol consumption questions (AUDIT-C): an effective brief screening test for problem drinking. Ambulatory Care Quality Improvement Project (ACQUIP). Alcohol Use Disorders Identification Test. Arch Intern Med. 1998;158(16):1789–95.

19. Espie CA, Kyle SD, Hames P, Gardani M et al. The Sleep Condition Indicator: a clinical screening tool to evaluate insomnia disorder. BMJ Open. 2014;4(3):e004183.

20. American Psychiatric Association. Diagnostic and Statistical Manual of Mental Disorders (DSM-5). Fifth edition. Washington DC: American Psychiatric Association 2013. 991p.

21. Schnurr PP, Spiro A, Vielhauer MJ, et al. Trauma in the Lives of Older Men: Findings from the Normative Aging Study. Journal of Clinical Geropsychology. 2002;8(3):175–87.

22. Bond FW, Hayes SC, Baer RA et al. Preliminary psychometric properties of the Acceptance and Action Questionnaire–II: a revised measure of psychological inflexibility and experiential avoidance. Behav Therapy. 2011;42(4): 676–688.

23. Sullivan T, Hansen P, Ombler F, et al. (2020) A new tool for creating personal and social EQ-5D-5L value sets, including valuing ‘dead’. Soc Sci Med. 2020; 246:112707.

24. Feng Y, Parkin D, Devlin NJ. Assessing the performance of the EQ-VAS in the NHS PROMs programme. Qual Life Res. 2014;23(3):977–89.

25. Brooks SK, Greenberg N. Non-deployment factors affecting psychological wellbeing in military personnel: literature review. J Ment Health. 2018;27(1):80–90.

26. Richardson A, Gurung G, Samaranayaka A, et al. Risk and protective factors for post-traumatic stress among New Zealand military personnel: A cross sectional study. PLoS One 2020;15(4):e0231460.

27. Boehmer TK, Boothe VL, Flanders WD et al. Health-related quality of life of U.S. military personnel: a population-based study. Mil Med. 2003;168(11):941–7.

28. Porter B, Long K, Rull RP, et al. Health Status of Gulf War and Era Veterans Serving in the US Military in 2000. J Occup Environ Med 2018;60(5):e261–e67.

29. Williamson V, Diehle J, Dunn R, et al. The impact of military service on health and well-being. Occup Med (Lond) 2019;69(1):64–70.

30. Diaz Santana MV, Eber S, Barth S, Cypel Y et al. Health-Related Quality of Life Among U.S. Veterans of Operation Enduring Freedom and Operation Iraqi Freedom-Results From a Population-Based Study. Mil Med. 2017;182(11):e1885–e91.

31. Newby JH, McCarroll JE, Ursano RJ, et al. Positive and negative consequences of a military deployment. Mil Med 2005;170(10):815–9.

32. McCarthy E, DeViva JC, Norman SB, et al. Self-assessed sleep quality partially mediates the relationship between PTSD symptoms and functioning and quality of life in U.S. Veterans: Results from the National Health and Resilience in Veterans Study. Psychol Trauma. 2019;11(8):869–76.

33. Wolgast M. What does the Acceptance and Action Questionnaire (AAQ-II) really measure? Behav Ther. 2014;45(6):831–839.

34. Hayes SC, Luoma JB, Bond FW, Masuda A, Lillis J. Acceptance and commitment therapy: model, processes and outcomes. Behav Res Ther. 2006;44(1):1–25.

35. Lang AJ, Schnurr PP, Jain S, He F, Walser RD, Bolton E, et al. Randomized controlled trial of acceptance and commitment therapy for distress and impairment in OEF/OIF/OND Veterans. Psychol Trauma. 2017;9(Suppl 1):74–84.

36. Troxel WM et al. Sleep in the Military: Promoting Healthy Sleep Among U.S. Servicemembers. RAND Corporation, 2015[cited 2020 September 4]. Available from: https://www.rand.org/content/dam/rand/pubs/research_reports/RR700/RR739/RAND_RR739.pdf.

37. Sweetman A et al. A step-by-step model for a brief behavioural treatment for insomnia in Australian general practice. Aust J Gen Pract. 2021;50(5):287–293.

