## Supplemental table for "Factors associated with self-reported health among New Zealand military Veterans: a cross-sectional study"

**Supplementary Table: Number of participants and EQ-VAS scores according to sample characteristics**

| Exposure variable | N(%) | | Mean (SD) |
|---|---|---|---|
| **All** | 1767 | | 74.4 (17.1) |
| **Age group (years)** | | | |
| 20-29 | 136 | (7.7) | 77.7 (14.4) |
| 30-39 | 328 | (18.6) | 77.0 (16.2) |
| 40-49 | 438 | (24.8) | 76.1 (14.6) |
| 50-59 | 350 | (19.8) | 74.8 (16.6) |
| 60-69 | 285 | (16.1) | 71.4 (19.8) |
| 70-79 | 175 | (9.9) | 69.4 (19.4) |
| 80+ | 50 | (2.8) | 66.2 (20.3) |
| Missing | 5 | (0.3) | 61.2 (22.1) |
| **Gender** | | | |
| Female | 220 | (12.5) | 70.3 (17.2) |
| Male | 1520 | (86.0) | 74.4 (17.2) |
| Missing | 27 | (1.5) | 75.0 (55.0) |
| **Ethnicity (prioritised)** | | | |
| NZ European | 1382 | (78.2) | 74.4 (17.1) |
| Māori | 245 | (13.9) | 75.6 (16.1) |
| Other | 140 | (7.9) | 71.5 (19.3) |
| **Service years** | | | |
| 0-9 | 339 | (19.2) | 69.9 (19.7) |
| 10-19 | 478 | (27.1) | 74.5 (17.9) |
| 20-29 | 530 | (30.0) | 75.3 (15.9) |
| 30-39 | 254 | (14.4) | 76.3 (14.6) |
| 40+ | 69 | (3.9) | 75.4 (16.4) |
| Missing | 97 | (5.5) | 78.8 (14.2) |
| **Deployment (ever)** | | | |
| No | 288 | (16.3) | 69.5 (19.9) |
| Yes | 1458 | (82.5) | 75.3 (16.4) |
| Missing | 21 | (1.2) | 75.6 (14.6) |
| **GHQ12 Score** | | | |
| 0-9 | 652 | (36.9) | 82.0 (12.6) |
| 10-19 | 972 | (55.0) | 72.3 (16.0) |
| 20-29 | 123 | (7.0) | 54.3 (20.3) |
| 30+ | 18 | (1.0) | 46.2 (24.4) |
| Missing | 2 | (0.1) | - |

*sub-scores not used in multivariable models.

Supplementary table contd.

| | | | |
|---|---|---|---|
| **Social support full score** | | | |
| 24-29 | 0 | 0 | - |
| 30-39 | 3 | (0.2) | 66.7 (15.3) |
| 40-49 | 15 | (0.8) | 44.5 (22.5) |
| 50-59 | 97 | (5.5) | 62.4 (21.8) |
| 60-69 | 351 | (19.9) | 69.0 (18.2) |
| 70-79 | 643 | (36.4) | 75.1 (15.0) |
| 80-89 | 409 | (23.1) | 78.0 (15.0) |
| 90-96 | 235 | (13.3) | 81.9 (13.9) |
| Missing | 14 | (0.8) | 58.1 (25.6) |
| **Social support sub-scores** | | | |
| **Attachment*** | | | |
| 4-7 | 51 | (3.5) | 61.4 (23.4) |
| 8-11 | 516 | (29.2) | 68.7 (18.0) |
| 12-16 | 1193 | (67.5) | 77.5 (15.5) |
| Missing | 7 | (0.4) | 61.0 (19.7) |
| **Social integration*** | | | |
| 4-7 | 20 | (1.1) | 56.2 (25.1) |
| 8-11 | 429 | (24.3) | 67.1 (19.4) |
| 12-16 | 1309 | (74.8) | 77.1 (15.2) |
| Missing | 9 | (0.5) | 58.7 (21.5) |
| **Reassurance of worth*** | | | |
| 4-7 | 37 | (2.1) | 59.1 (19.5) |
| 8-11 | 504 | (28.5) | 69.0 (19.1) |
| 12-16 | 1215 | (68.8) | 77.2 (15.2) |
| Missing | 11 | (0.6) | 62.0 (24.9) |
| **Reliable Alliance*** | | | |
| 4-7 | 20 | (1.1) | 49.6 (22.2) |
| 8-11 | 251 | (14.2) | 66.9 (20.1) |
| 12-16 | 1489 | (84.3) | 76.0 (15.9) |
| Missing | 7 | (0.4) | 61.0 (19.7) |
| **Guidance sub-score*** | | | |
| 4-7 | 36 | (2.0) | 59.2 (22.2) |
| 8-11 | 387 | (21.9) | 68.7 (18.9) |
| 12-16 | 1337 | (75.7) | 76.6 (15.7) |
| Missing | 7 | (0.4) | 54.6 (21.6) |
| **Opportunity for nurturance*** | | | |
| 4-7 | 29 | (1.6) | 69.3 (18.9) |
| 8-11 | 413 | (23.4) | 71.9 (18.8) |
| 12-16 | 1316 | (74.5) | 75.4 (16.4) |
| Missing | 9 | (0.5) | 58.7 (21.5) |

Supplementary table, contd.

| | | | |
|---|---|---|---|
| **Psychological flexibility score (AAQii score).** | | | |
| 10-19 | 5 | (0.3) | 23.2 (13.8) |
| 20-29 | 40 | (2.3) | 49.1 (22.1) |
| 30-39 | 147 | (8.3) | 63.0 (18.6) |
| 40-49 | 43 | (24.7) | 68.5 (16.8) |
| 50-59 | 664 | (37.6) | 77.1 (13.9) |
| 60-70 | 457 | (25.9) | 82.5 (13.5) |
| Missing | 1 | (1.0) | 69.6 (20.5) |
| **Sleep Condition Indicator (SCI score** | | | |
| 0 to <2 | 67 | (3.8) | 51.2 (22.3) |
| 2 to <4 | 264 | (14.9) | 62.2 (19.5) |
| 4 to <6 | 646 | (36.6) | 74.3 (14.6) |
| 6 to <8 | 384 | (21.7) | 77.6 (14.2) |
| 8 to 10 | 386 | (21.8) | 83.5 (12.2) |
| Missing | 20 | (1.1) | 77.3 (12.8) |
| **AUDIT-C score** | | | |
| Non-hazardous | 776 | (43.9) | 74.0 (17.4) |
| Hazardous | 915 | (51.8) | 74.6 (16.9) |
| Missing | 76 | (4.3) | 75.0 (17.0) |
| **Brief Trauma Questionnaire (DSM-IV criteria)** | | | |
| Not exposed | 544 | (30.8) | 78.3 (15.1) |
| Exposed | 1210 | (68.5) | 72.5 (17.8) |
| Missing | 13 | (0.7) | 80.6 (8.1) |
